# Death Registration coverage in India: Results from nationally representative survey

**DOI:** 10.1101/2022.08.25.22279168

**Authors:** Nandita Saikia, Krishna Kumar, Bhaswati Das

## Abstract

**Objective:** To investigate the disparity and predictors of death registration in India.

**Methods:** We used National Family Health Survey (NFHS-2019-21) data. Based on eligible household members’ reports, we estimated death registration coverage among 84,390 deaths in all age groups across the country. We did multilevel binary logistic regression to examine demographic and socio-demographic predictor variables of death registration at state, district, and individual levels. We used GIS software for spatial mapping of the level of death registration at the district level, disaggregated by sex.

**Findings:** The death registration at the national level is 71%. We found that out of 707 districts in 2019, 122 and 53 districts recorded death registration level below 40 percent among females and males, respectively. There was a considerable difference in the death registration level by sex (male-74% and female-66%). Death registration level was higher in urban areas compared to rural areas (83% vs. 66%). We found death registration level was higher among households with BPL cards (72%), bank accounts (71%) and covered with health insurance (77%). Females, rural populations, people from disadvantaged castes, poorest wealth quintile, Muslims, and not having BPL cards have a lower likelihood of death registration in India. District-level predictors were not statistically significant in the model.

**Conclusion:** Demographic and socioeconomic characteristics of the deceased are significantly associated with their death registration. We suggest periodic awareness programs on death registration procedures and facilitating easy access to death registration offices in lower performing districts and areas among the marginalised population groups.

## Introduction

Accurate mortality statistics are an indispensable tool for healthcare utilization planning, resource allocation, and policymaking.^1^ Mortality data by age, sex and cause of death and place of residence is essential for health officials and decision-makers to identify health threats and high-risk populations.^2^ Sustainable Development Goal 3 states, “*ensure healthy living and promote well-being for all ages by 2030*”. The goal primarily includes targets to reduce maternal and child death rates and other premature deaths from non-communicable diseases.^3^ Measurement of these targets is difficult without continuous and complete death registration, for measuring SDG targets on under-five mortality and other premature deaths.^4^ The SDG 19.17 states tracking progress in birth and death registration for monitoring statistical capacity.^3^ The WHO SCORE report also suggests strengthening overall health data systems, improving their death registration level and collecting better quality data to address inequality.^5^

There is no alternative of the Civil Registration System (CRS) to produce high-quality mortality statistics. Yet, in many low and middle-income countries, CRS is still deficient. With the availability of Demographic Health Surveys (DHS), a high-quality data for children’s deaths are generated, however, adult and old age death statistics still suffer serious data quality issues. In addition, DHS cannot generate mortality estimates for vulnerable populations due to sample size restrictions. Thus, estimation of mortality is challenging, and quality of information is unsatisfactory in many lower and middle-income countries since death registration in these countries is not universal.^6,7^ Notably, most people in Africa and Asia do not register birth and death, which has huge implications for legal identities and officials’ statistics.^7,8^ Accurate death registration data also help the judiciary and individual to resolve inheritance issues fairly.^2,9^ The broke out of the Covid pandemic reiterated the importance of accurate counting of deceased and facilitating monitoring of the epidemic.

In India, the Registration of Births and Deaths Act (RBD Act) mandates the registration of all births and deaths within 21 days.^9^ The number of registered death has gone up from 7.64 million in 2019 to 8.12 million in 2020, i.e. an increase of 6.2%. (Office of Registrar General of India^9^), however, there is a wide disparity in death registration by states.

Mortality data generated from civil registration systems have been used less due to high underreporting of death in many states^9^. Previous studies in other countries showed death registration was higher among educated mothers, major ethnic groups and non-poor households.^8,10^ However, there are limited studies in India examining intra-district variation in death registration. To our knowledge, there is no published research article that shows a deceased’s death registration by their demographic and socioeconomic characteristics. The recently available data of the National Family Health Survey (2019-21) gives a unique opportunity to examine death registration by socioeconomic characteristics. Death registration by socioeconomic factors helps to identify the target population for awareness of death registration. This study aims to investigate the district (administrative units) level disparity and socioeconomic disparity in death registration in India.

## Methods

### Data source

We used data from the Indian DHS, popularly known as National Family Health Survey, 2019-21 (NFHS-5), conducted under the Ministry of Health and Family Welfare (MoHFW) and the International Institute of Population Sciences (IIPS), Mumbai. It is a nationally representative survey and gives reliable information on household populations, housing characteristics, fertility, family planning, maternal and child health, stillbirth, infant and child mortality, death registration, nutrition, morbidity, including adult health issues for 28 states, 8 Union Territory (UT) and 707 districts. The survey collects information from a total of 636,699 households with a response rate of 98 percent. In the selected households, 724,115 women and 101,839 men were interviewed. We included a total sample of 84,390 individuals who died in last five years of survey in the final analysis. The details of the sample survey is given elsewhere.^11^

### Data collection

The NFHS provides information on the number of deaths in a household since 2016. In the survey, a question on death registration was also asked from family members as “Death registered with the civil authority”.^11^ A dependent variable “death registration with civil authority” was created as binary variable with 1 equals to individual’s death registered, otherwise 0.

We considered the demographic and socioeconomic variables of the deceased person. We categorised deceased’s age as 12-34, 35-44, 45-54, 55-64, 65-69, and 70 and above years, sex of the deceased (male or female). Other demographic and socioeconomic characteristics include the place of residence (urban or rural), region (north, north-east, south, central, east, west), highest level of education completed by the household members (illiterate, primary, secondary and higher), religion of the head of household (Hindus, Muslims and other), caste of the head of household (Scheduled Caste (SCs), Scheduled Tribes (STs), Other Backward Caste (OBC) and Other), wealth quintile (poorest, poorer, middle, richer, richest), and types of family (nuclear or non-nuclear). Furthermore, we included welfare variables as household has a BPL card (yes or no), household has a bank account (yes or no) and the usual members of a household covered by health insurance (yes or no).

In addition, we considered the district level variables such as proportion of ST population, proportion of household where at least one household members completed secondary education, mean household size, proportional of household having bank account, proportional of household covered by health insurance, proportional of household live in urban areas, proportional of institutional birth in a district.

### Data analysis

First, we estimated death registration at the district level. We also estimated death registration by socioeconomic characteristics of the deceased. We performed a chi-square test to examine the association of demographic, socioeconomic and welfare variables associated with the deceased’s death registration. We then applied multilevel binary logistic regression models with random intercept and fixed slope to calculate the Adjusted odds ratio (AOR) at three levels (level 1: individual; level 2: district; level 3: state) with 95 percent of confidence interval (CI). We used the survey weights to adjust the design of the study. We considered odds ratios significant; when the p-value was, lower than 0.05. Multilevel analysis generates variance at each level, providing the technical advantage of assessing unobserved effects at each level.

We used Akaike Information Criteria (AIC) and log-likelihood for model comparison. Besides, we checked multicollinearity using the Variance Inflation Factor (VIF). We found no collinearity among the included independent variables (mean VIF = 1.43). Intra Class Correlation (ICC) was also estimated to show the percentage variance explained at the district and state levels. All analysis was performed using R (version 4.0.2). Further, we also mapped the district-wise proportion of registered death using the Geographic Information System (GIS).

## Results

Appendix 1 presents descriptive statistics of outcome and predictor variables included in this study. We found that 71 percent of the deceased in our sample was registered with a civil authority, as reported by the household members.

Fig 1 shows a glaring gender difference in death registration. Death registration among males is much higher than that of females across Indian districts. Secondly, fig 1 shows a wide geographical disparity in death registration. Out of 707 districts in 2019, 122 districts showed female death registration level below 40 percent and 251 districts recorded death registration level above 80 percent. In Bihar, out of 38 districts, 37 districts showed death registration level below 60 percent. Further, all the districts of Jharkhand, 18 districts of Arunachal Pradesh and 70 districts of UP showed lower than 60 percent death registration level. While the Mumbai district of Maharashtra has as high as 100% coverage of deaths, Kurung Kumey district of Arunachal Pradesh has the lowest registration of 5.39%. Besides, Fig 1 also shows death registration level among male by India’s districts during 2019-2021. We found, 53 districts recorded male death registration level below 40 percent whereas 354 districts reported death registration above 80 percent. Moreover, 17 districts namely, Mumbai, Kannur, Rajkot, Thrissur, Kancheepuram, Valsad etc. reported 100% coverage of deaths. On the other hand, Kurung Kumey (10.01%) and Upper Subansiri (10.24%) district of Arunachal Pradesh has the lowest registration of death. We found death registration level vary greatly by districts and sex.

**Fig 1.**
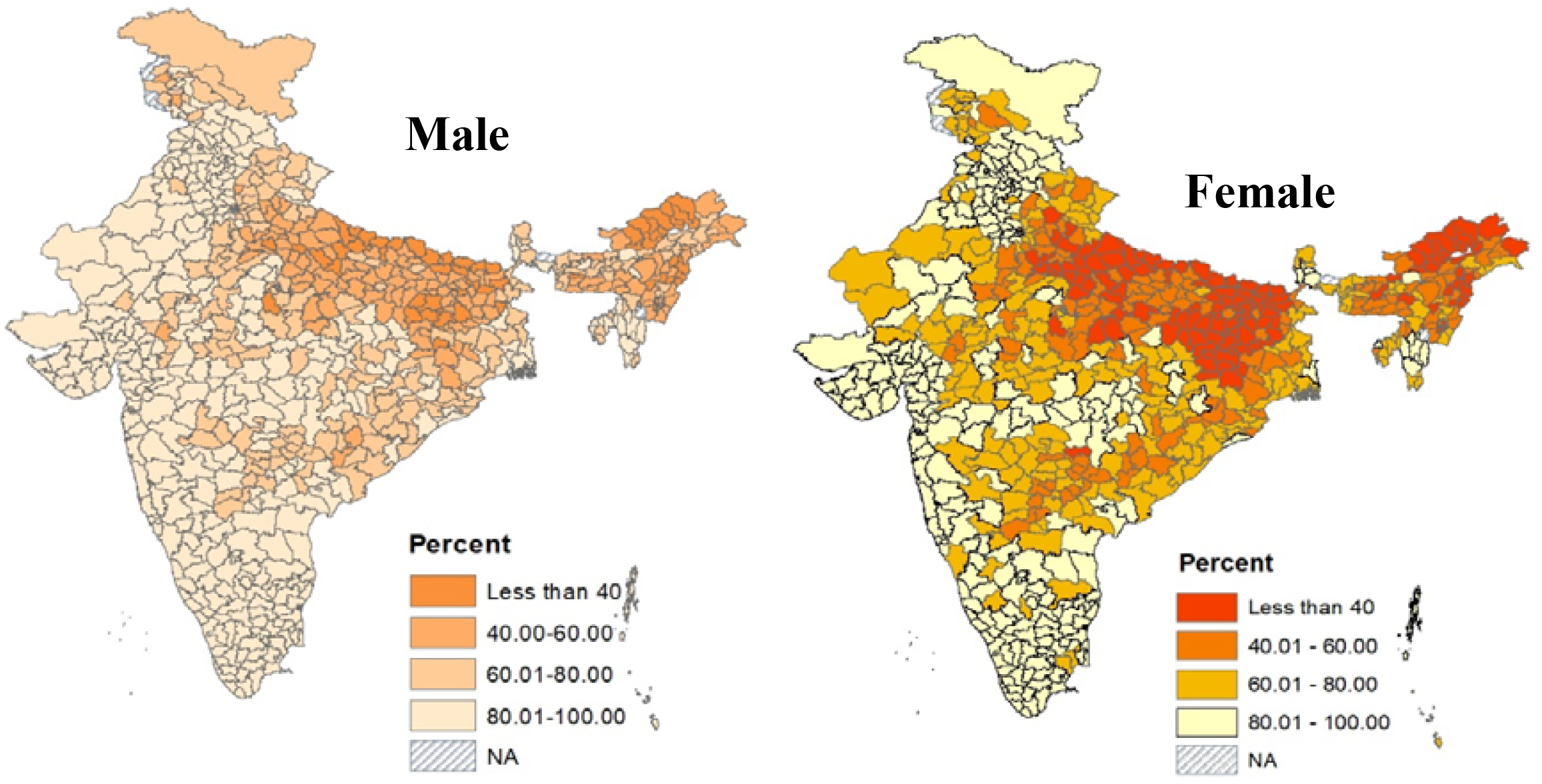
Level of death registration by male and female in the districts of India, 2019-2021. Source: Authors created this map using GIS

Fig 2 presents the death registration coverage by the deceased’s own or household characteristics. We found a deceased’s age is significantly associated with his or her death registration. Death registration level was found to be higher at an older age (64% at 12-34 years and 74% at age 70 and above). There was a considerable difference in death registration levels by sex (male-74% and female-66%). Death registration level was higher in urban areas compared to rural areas (83% vs 66%). We found household members’ highest level of education, wealth status of the household, religion and caste of the head of household, and types of the family were found significantly associated with death registration. We observed a lower death registration level among households where any member of the family was illiterate (63%) and completed primary education (71%). A higher death registration level was found in the richest (87%) and richer (81%) households as compared to the poorest households (52%). Death registration level was lower among Muslims (65%) as compared to Hindus (71%). Besides, the death registration level was higher among other castes (77%), compared to STs (67%). We found there is a marginal difference in death registration level by types of family (nuclear family-70% and non-nuclear family-71%). Death registration level was higher among the households, which has a BPL card (72%), bank account (71%) and covered with health insurance (77%).

**Fig 2.**
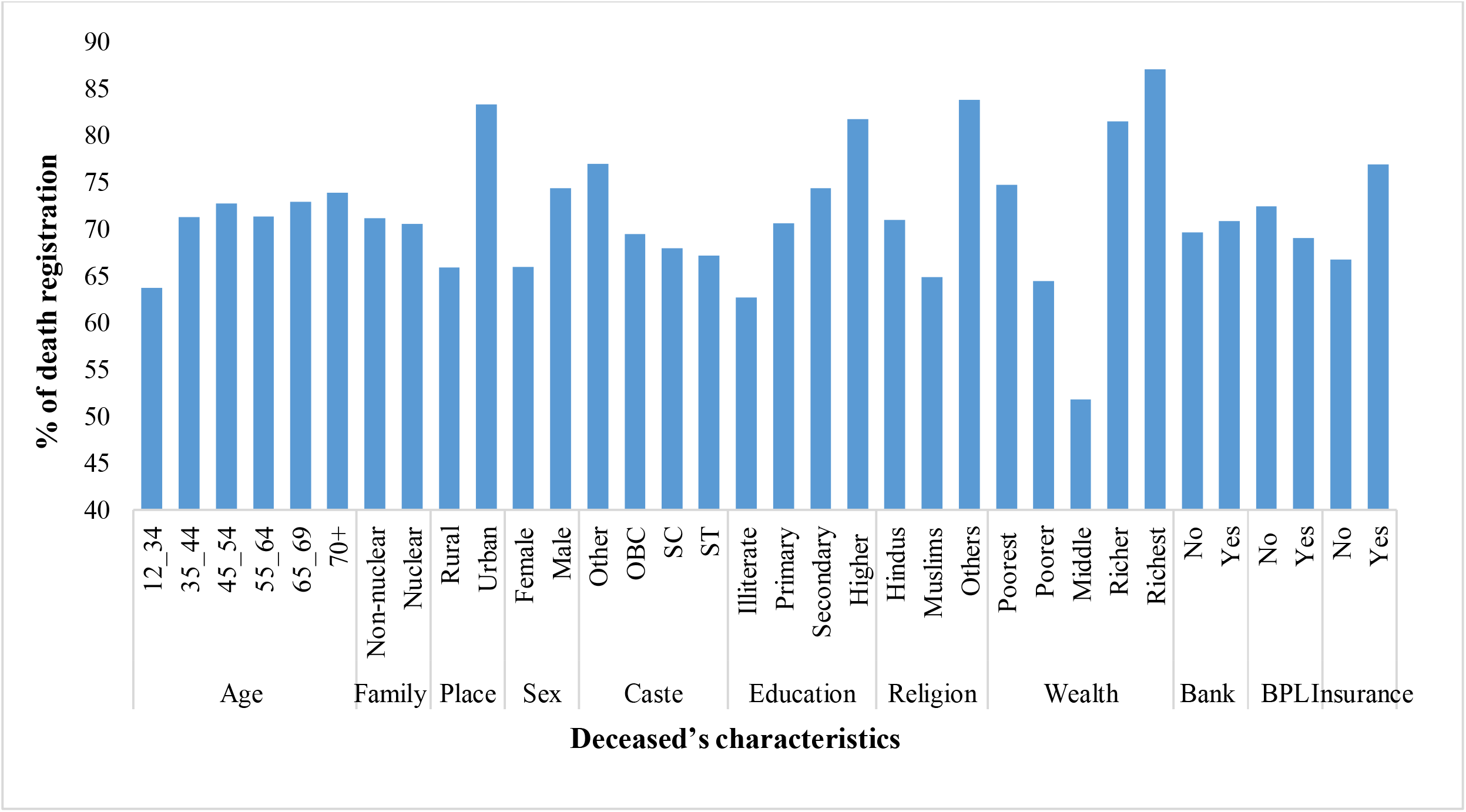
Death registration level by demographic and socioeconomic characteristics, India, 2019-21. Source: Authors’ calculations

Table 1 shows the result of multilevel binary logistic regression of demographic, socioeconomic and welfare factors. The random variance of 0.53, 0.16 and 3.29 at the state, district and individual level, respectively. In addition, an ICC value of 0.13 at the state level showed that 13 percent of the total variation in death registration level is explained by between state-level differences while the remaining 87 percent lies within states. We found the ICC value at the district level was 0.04 indicating 4 percent of total variation in death registration level lies between districts. The finding highlights a need for adequate programmes to improve death reporting at the lower administrative level.

**Table 1.** Results of multilevel binary logistic regression of death registration by demographic and socioeconomics characteristics, India, 2019-21.

| Fixed effect parameter | AOR (95%) CI |
| --- | --- |
| Intercept | 4.39 |
| **Age of the deceased** |  |
| 12_34 | Reference |
| 35-44 | 1.19 (1.12 to 1.26) |
| 45-54 | 1.15 (1.08 to 1.22) |
| 55-64 | 1.13 (1.06 to 1.20) |
| 65-69 | 1.22 (1.13 to 1.32) |
| 70+ | 1.22 (1.13 to 1.32) |
| **Sex of the deceased** |  |
| Male | Reference |
| Female | 0.61 (0.58 to 0.63) |
| **Place of residence** |  |
| Urban | Reference |
| Rural | 0.76 (0.72 to 0.81) |
| **Region** |  |
| North | Reference |
| North-East | 0.35 (0.17 to 1.72) |
| South | 1.49 (0.69 to 3.20) |
| Central | 0.40 (0.15 to 1.06) |
| East | 0.28 (0.12 to 0.69) |
| West | 2.89 (1.13 to 7.39) |
| **highest education level completed** |  |
| Illiterate | Reference |
| Primary | 1.08 (1.02 to 1.15) |
| Secondary | 1.26 (1.21 to 1.31) |
| Higher | 1.57 (1.45 to 1.70) |
| **Wealth Quintile** |  |
| Poorest | Reference |
| Poorer | 1.26 (1.19 to 1.33) |
| Middle | 1.54 (1.45 to 1.63) |
| Richer | 1.82 (1.72 to 1.93) |
| Richest | 2.34 (2.16 to 2.53) |
| **Religion** |  |
| Hindus | Reference |
| Muslims | 0.79 (0.73 to 0.85) |
| Others | 0.97 (0.88 to 1.07) |
| **Caste** |  |
| SC | Reference |
| ST | 0.85 (0.80 to 0.90) |
| OBC | 1.04 (1.00 to 1.08) |
| Other | 1.14 (1.07 to 1.21) |
| **Type of family** |  |
| Nuclear | Reference |
| Non-nuclear | 1.02 (0.98 to 1.06) |
| **Have a BPL Card** |  |
| No | Reference |
| Yes | 1.07 (1.03 to 1.12) |
| **District level demographic variable** |  |
| Proportion of ST in a district | 0.90 (0.74 to 1.10) |
| Percent secondary educated | 0.78 (0.54 to 1.13) |
| Mean household size | 1.01 (0.95 to 1.07) |
| **District level welfare variable** |  |
| Proportion having bank account | 2.14 (0.40 to 11.54) |
| Proportion having health insurance | 0.99 (0.83 to 1.18) |
| Proportion reside in urban areas | 1.15 (0.68 to 1.95) |
| Proportion of institutional birth | 1.04 (0.75 to 1.45) |
| **Random effect parameter** |  |
| State level | 0.54 (0.74) |
| District level | 0.16 (0.40) |
| Individual level | 3.29 (1.81) |
| **ICC** |  |
| ICC at state level | 0.14 |
| ICC at district level | 0.04 |
| **AIC** | 77429.00 |
| **Log Likelihood** | -38678.40 |
| **VIF** | 1.43 |
| **Number of state** | 36 |
| **Number of district** | 707 |
| **Number of observation** | 77697 |

Source- Authors' calculations
Note: In random effect parameter, coefficients (standard errors) are shown.

We found as compared with deceased aged 12-34 years, deceased among the age group 35-44 years (AOR:1.19; 95% CI: 1.12 to 1.26), 45-54 years (AOR:1.15; 95% CI: 1.08 to 1.22) and 70 and above years (AOR:1.22; 95% CI: 1.13 to 1.32) have a higher likelihood of death registration. Interestingly, the odds of female death registration was found to be lower (AOR: 0.61; 95% CI: 0.58 to 0.63) as compared with male. We found the place of residence of a deceased person is significantly associated with his or her death registration. As compared with death registration in urban areas, rural areas showed a lower (AOR: 0.76; 95% CI: 0.72 to 0.81) likelihood of death registration. Similarly, we found that India’s region is highly associated with death registration practices. As compared with north region, northeast (AOR: 0.35; 95% CI: 0.17 to 0.72), central (AOR: 0.40; 95% CI: 0.15 to 1.06) and east region (AOR: 0.28; 95% CI: 0.12 to 0.69) showed a lower odds of death registration. On the other hand, the south (AOR=1.49; 95% CI: 0.69 to 3.20) and the west region (AOR: 2.89; 95% CI: 1.13 to 7.39) showed a higher likelihood for death registration as compared with the north region.

We found odds of death registration are higher among secondary (AOR: 1.26; 95% CI: 1.21 to 1.31) and higher educated household members (AOR: 1.57; 95% CI: 1.45 to 1.70). A deceased household’s wealth status is also significantly associated with death registration. As compared with the poorest household, richer (AOR: 1.82; 95% CI: 1.72 to 1.93) and richest (AOR: 2.34; 95% CI: 2.16 to 2.53) households showed a higher likelihood for death registration. Besides, this study showed that religion and caste are significantly associated with death registration. We found a lower odd of death registration among Muslims as compared with Hindus (AOR: 0.79; 95% CI: 0.73 to 0.85). Furthermore, a lower likelihood for death registration was observed among STs (AOR: 0.85; 95% CI: 0.80 to 0.90), as compared to SC households. In addition, we found a higher odd of death registration among households which was provided a BPL card (AOR: 1.07; 95% CI: 1.03 to 1.12).

The study also found that living in a district with a higher proportion of STs households decreases the odds of deceased’s death registration (AOR: 0.90; 95% CI: 0.74 to 1.10). However, the result was not significant. We did not find any significant association between death registration and the other district-level variable.

## Discussion

The wide disparity in access to death registration is unequal socioeconomic development, lack of awareness and lack of facilities. The level of death registration is about 71% at the national level but varies from 5 percent to 100 percent at the district level. South and West regions showed a higher level of death registration as compared with the north region. The majority of western states and southern states are better performing states in terms of SDGs index^12^,which could result in a higher death registration level.

A clear gender disparity in death registration is observed with a lower level of registration for females. A sizeable sex difference in death registration could be attributed to a lower proportion of females employed in the formal sector; therefore, a lower incentive is associated with female death registration.^13,14^ In addition, a global scenario showed that female has a longer life expectancy than male, and India is not an exception to this fact, which may mean that there is no one to register a wife’s death after the death of the husband in a single household.^15^ A higher proportion of accidental deaths among males (which are usually the subject of a police investigation) may lead to higher odds of death registration among males.^16,17^

We found that death registration is lower among all the disadvantaged groups, such as among women, rural residents, economically poor and deprived caste groups. An earlier study found that socioeconomic development is a major determinant of healthcare utilisation programmes, strategies and activities, including reporting the death to a civil authority. Socioeconomic development led to achieving other factors related to strengthening death registration services.^18^ This study showed higher odds of death registration among older people. The higher registration level at an older age could be associated with inheritance, pension claims, and insurance.^17,19^

The place of residence of the deceased is significantly associated with his or her death registration. In rural areas, the majority of adults are employed in informal sectors such as farming, cultivation, construction work and fishing which seldom provides social security such as a family pension. The absence of motivation, few incentives and poor access to death registration services lead to a lower registration level in rural areas as compared to urban areas.^13,20^ The level of education completed by household members of the deceased showed a significant association with death registration. Previous studies also documented knowledge-related barriers limit the coverage of death registration in low and middle-income countries.^18,21^

Deceased from economically well-off households have a higher level of death registration. A previous study showed wealthier households take patients to hospitals and consequently afford the costs incurred in death registration.^19^ Household income was significantly associated with death registration.^18^

There is lower reporting of death among Muslims and STs households. A previous study showed cultural beliefs and traditional practices are the reasons for the delay in death registration in Indonesia.^22^ In China, given their own culture and beliefs, most people prefer to die at home and death information is not reported to civil authorities.^23^ Under-reporting of deaths among Muslims and STs may be due to some cultural belief, which needs further investigation.

BPL cardholder households showed a higher odd for death registration. A family member of the deceased who was a BPL cardholder may seek financial assistance due to premature death. Therefore, there is a higher possibility of death registration. Under National Family Benefit Schemes, a lump sum family benefit of rupees 10000 is provided to the bereaved households in case of death of a primary breadwinner. Only those families who hold BPL cards are eligible for this scheme.^24^

Our findings suggest that reporting variation is explained by mainly individual-level variables (about 96 percent). Living in a district where a higher proportion of secondary educated people reside would definitely result in a high level of awareness regarding the death registration process. However, most of the district-level variables are not statistically significant in our study.

While our findings indicate that poorer socioeconomic background is associated with a lower registration level, earlier studies showed that contextual factors lead to insufficient death registration. For example, the district has several registration offices, and the distance to the registration centre can affect the death registration.^25,26^ A detailed study in Bihar recently found that people faced challenges in reporting birth and death due to poor delivery of services at the registration centres, higher indirect opportunity cost, and demand of bribes by the CRS staff for providing certificates. There was also a lack of adequate investment, a shortage of dedicated staff, and limited computer and internet services at the registration centres.^27^

This study has some limitations. First, death registration data are captured as respondents replied; the interviewer did not check for the death certificate. Therefore, this study’s findings may be over-reported in case a respondent misunderstood the deceased’s medical record as the death certificate. Secondly, part of the data collection was done in the post-Covid period. Due to the government compensation scheme for Covid-related deaths, there is a chance of higher coverage of deaths in the system than usual time. Thirdly, NFHS is a cross-sectional survey; therefore, we do not claim causality between the death registration and socioeconomic variables. Despite the above limitations, this study is a novel attempt to fill the gap in the existing literature on death registration in India.

## Conclusion

Many countries in South Asia missed more than one-third of death records.^2^ We found that age, sex, education, religion, caste, and wealth status are significant covariates of death registration. Besides, place of residence and region of residence are crucial to getting death registered. We suggest periodic awareness programs among socially disadvantaged populations and demographically backwards areas. Creating demand for death registration by providing financial assistance for performing funeral rites, education loans to orphans, and social security to the deceased’s family member after reporting of death to a civil authority can help to increase the death registration level in India.

## Data Availability

Data are publically available on the DHS website

https://dhsprogram.com/data/dataset_admin/login_main.cfm

---

**Appendix 1:** Descriptive statistics of the sample of the study population, India, NFHS, 2019-21.

|  | Percent | S.E | Confidence Interval |  |
| --- | --- | --- | --- | --- |
|  |  |  | Lower Limit | Upper Limit |
| **Death registered with a civil authority** |  |  |  |  |
| No | 29.21 | 0.00 | 28.81 | 29.62 |
| Yes | 70.79 | 0.00 | 70.38 | 71.19 |
| **Age of the deceased** |  |  |  |  |
| 12_34 | 14.91 | 0.00 | 14.85 | 14.97 |
| 35_44 | 22.34 | 0.00 | 22.28 | 22.41 |
| 45_54 | 24.36 | 0.00 | 24.29 | 24.42 |
| 55_64 | 21.21 | 0.00 | 21.15 | 21.27 |
| 65_69 | 7.70 | 0.00 | 7.66 | 7.74 |
| 70+ | 9.48 | 0.00 | 9.44 | 9.53 |
| **Sex of the deceased** |  |  |  |  |
| Male | 58.04 | 0.00 | 57.82 | 58.25 |
| Female | 41.97 | 0.00 | 41.75 | 42.18 |
| **Place of residence** |  |  |  |  |
| Urban | 33.18 | 0.00 | 33.10 | 33.26 |
| Rural | 66.82 | 0.00 | 66.74 | 66.90 |
| **Region** |  |  |  |  |
| North | 12.92 | 0.00 | 12.88 | 12.96 |
| North-East | 3.73 | 0.00 | 3.71 | 3.74 |
| South | 23.57 | 0.00 | 23.50 | 23.64 |
| Central | 22.53 | 0.00 | 22.47 | 22.59 |
| East | 22.52 | 0.00 | 22.45 | 22.59 |
| West | 14.73 | 0.00 | 14.66 | 14.80 |
| **Highest education level completed** |  |  |  |  |
| Illiterate | 28.48 | 0.00 | 28.41 | 28.54 |
| Primary | 18.62 | 0.00 | 18.56 | 18.68 |
| Secondary | 41.98 | 0.00 | 41.91 | 42.06 |
| Higher | 10.92 | 0.00 | 10.86 | 10.97 |
| **Wealth Quintile** |  |  |  |  |
| Poorest | 20.78 | 0.00 | 20.72 | 20.84 |
| Poorer | 19.99 | 0.00 | 19.93 | 20.05 |
| Middle | 19.96 | 0.00 | 19.90 | 20.03 |
| Richer | 19.62 | 0.00 | 19.56 | 19.69 |
| Richest | 19.64 | 0.00 | 19.57 | 19.71 |
| **Religion** |  |  |  |  |
| Hindus | 81.90 | 0.00 | 81.84 | 81.97 |
| Muslims | 12.40 | 0.00 | 12.34 | 12.45 |
| Others | 5.70 | 0.00 | 5.66 | 5.73 |
| **Caste** |  |  |  |  |
| SC | 22.93 | 0.00 | 22.86 | 23.00 |
| ST | 10.07 | 0.00 | 10.03 | 10.11 |
| OBC | 44.06 | 0.00 | 43.98 | 44.14 |
| Other | 22.94 | 0.00 | 22.86 | 23.01 |
| **Family type** |  |  |  |  |
| Nuclear Family | 58.21 | 0.00 | 58.13 | 58.28 |
| Non-nuclear Family | 41.79 | 0.00 | 41.72 | 41.87 |
| **Has a BPL Card** |  |  |  |  |
| No | 54.83 | 0.00 | 54.76 | 54.91 |
| Yes | 45.17 | 0.00 | 45.09 | 45.24 |
| **Has a bank account** |  |  |  |  |
| No | 4.26 | 0.00 | 4.22 | 4.29 |
| Yes | 95.74 | 0.00 | 95.71 | 95.78 |
| **Cover by health insurance** |  |  |  |  |
| No | 58.78 | 0.00 | 58.71 | 58.86 |
| Yes | 41.22 | 0.00 | 41.14 | 41.29 |

Source: Authors' calculation

